# Universality in COVID-19 spread in view of the Gompertz function

**DOI:** 10.1101/2020.06.18.20135210

**Authors:** Akira Ohnishi, Yusuke Namekawa, Tokuro Fukui

**Author notes:** Report number: YITP-20-82.

## Abstract

We demonstrate that universal scaling behavior is observed in the current coronavirus (SARS-CoV-2) spread, the COVID-19 pandemic, in various countries. We analyze the numbers of infected people who tested positive (cases) in selected eleven countries (Japan, USA, Russia, Brazil, China, Italy, Indonesia, Spain, South Korea, UK, and Sweden). By using the double exponential function called the Gompertz function, *f_G_*(*x*) = exp(−*e*^−^*^x^*), the number of cases is well described as *N*(*t*) = *N*_0_*f_G_*(γ(*t* − *t*_0_)), where *N*_0_, 7 and *t*_0_ are the final number of cases, the damping rate of the infection probability and the peak time of the daily number of new cases, *dN*(*t*)/*dt*, respectively. The scaled data of cases in most of the analyzed countries are found to collapse onto a common scaling function *f_G_*(*x*) with *x* = γ(*t* − *t*_0_) being the scaling variable in the range of *f_G_*(*x*) ± 0.05. The recently proposed indicator so-called the *K* value, the increasing rate of cases in one week, is also found to show universal behavior. The mechanism for the Gompertz function to appear is discussed from the time dependence of the produced pion numbers in nucleus-nucleus collisions, which is also found to be described by the Gompertz function.

## 1. Introduction

The COVID-19 pandemic is the worst disease spread in this century. As of May 20, 2020, over 4 million people have tested positive in the world, and the number of cases *N*(*t*), the number of infected people who tested positive at the time *t*, is still increasing rapidly. In order to control the spread of infection, it is desired to understand the diffusion mechanism of COVID-19.

Recently, a double exponential function called the Gompertz function is found to catch the features of *N*(*t*) [1-20]. The Gompertz function appears when the infection probability per infected people exponentially decreases as a function of time. With the Gompertz function, the daily number of new cases *dN*(*t*)/*dt*, daily increase of infected people who tested positive, shows asymmetric time-profile rather than the symmetric one found in the prediction of the Susceptible-Infected (SI) model [21], one of the standard models of the spread of infection. The Gompertz function was proposed by B. Gompertz in 1825 to discuss the life contingencies [22]. It is interesting to find that the Gompertz function also appears as the number of tumors [23] and the number of detected bugs in a software [24], as well as particle multiplicities at high energies [25].

The exponential decrease of the infection probability is also important to deduce when the restrictions can be relaxed. For example, Nakano and Ikeda [26] found that the number of cases is well characterized by the newly proposed indicator *K*, which represents the increasing rate of cases in one week. The indicator *K* takes a value between zero and unity, is not affected by the weekly schedule of the test, and is found to decrease almost linearly as a function of time in the region 0.25 < *K* < 0.9 provided that there is only a single outbreak affecting the infection. In order to understand the linearly decreasing behavior of *K*, Nakano and Ikeda proposed the “constant damping hypothesis” in their paper [26], and Akiyama proved that the hypothesis of exponential decrease in discrete time shows the Gompertz curve in continuous time [27], independently of other works [1–20]. Since the indicator *K* is expected to be useful to predict the date when the restrictions can be relaxed as *K*(*t*) ≃ 0.05, it would be valuable to analyze its solution, the Gompertz function, in more detail.

In this article, we analyze the number of cases by using the Gompertz function. We examine that *N*(*t*) in one outbreak is well described by the Gompertz function in several countries. Then with the time shift and the scale transformation of time and *N*(*t*), the data are found to show universal behavior; they are on one-curve described by the basic Gompertz function, exp(−*e*^−^*^x^*). The newly proposed indicator *K* [26] also shows universality. We further discuss that the number of produced pions in nucleus-nucleus collisions is described by the Gompertz function. This similarity may be helpful to understand the mechanism of COVID-19 spread.

This article is organized as follows. In Sec. 2, we give a brief review of the Gompertz function and its relevance to the disease spread. In Sec. 3, we show the comparison of the number of cases and the Gompertz function fitting results. We demonstrate that the numbers of cases in many countries show universal scaling behavior. In Sec. 4, we show that the number of pions in nuclear collisions is well described by the Gompertz function, and we deduce the mechanism to produce the time-dependence described by the Gompertz function. In Sec. 5, we summarize our work.

## 2. Gompertz function, indicator *K* and scaling variables

When the infection probability *K*(*t*) is given as a function of time, the evolution equation for the number of infected people *N*(*t*) and its solution are given as

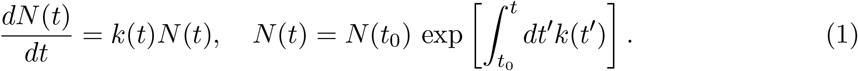

Adopting an exponentially decreasing function as *K*(*t*), the solution is found to be

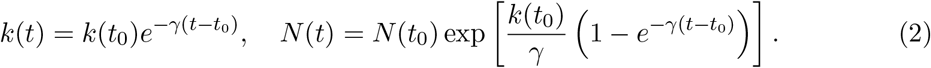

By choosing the reference time *t*_0_ so that *k*(*t*_0_) = γ, the solution is given as

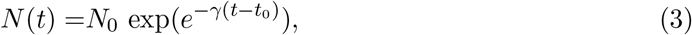

where *N*_0_ = *N*(*t*_0_) exp(*k*(*t*_0_)/γ)= *eN*(*t*_0_) is the asymptotic value of *N*(*t*), *N*_0_ = lim_t→∞_ *N*(*t*). Here, *N*_0_ can be interpreted as “terminal velocity”, since the evolution equation (1) has an analogy to the equation of motion of particle feeling a viscous resistance, where its coefficient is not constant but exponentially decreasing.

The double exponential function appearing in Eq. (3) is called the Gompertz function [22],

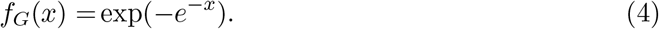

By using the Gompertz function, *N*(*t*) is given as

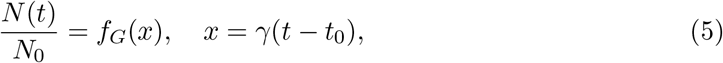

where *x* is the scaling variable for *N*(*t*).

One of the characteristic features of the Gompertz function is the asymmetry of its derivative in the early (*x*< 0) and late (*x*> 0) stages. In Fig. 1, we show the Gompertz function *f_G_*(*x*) and its derivative,

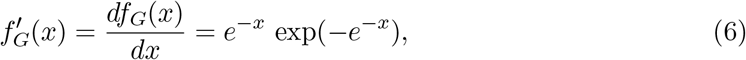

as functions of *x*. The derivative takes the maximum at *x* = 0, and the asymmetry in the negative and positive *x* region is clearly seen.

**Fig. 1.**
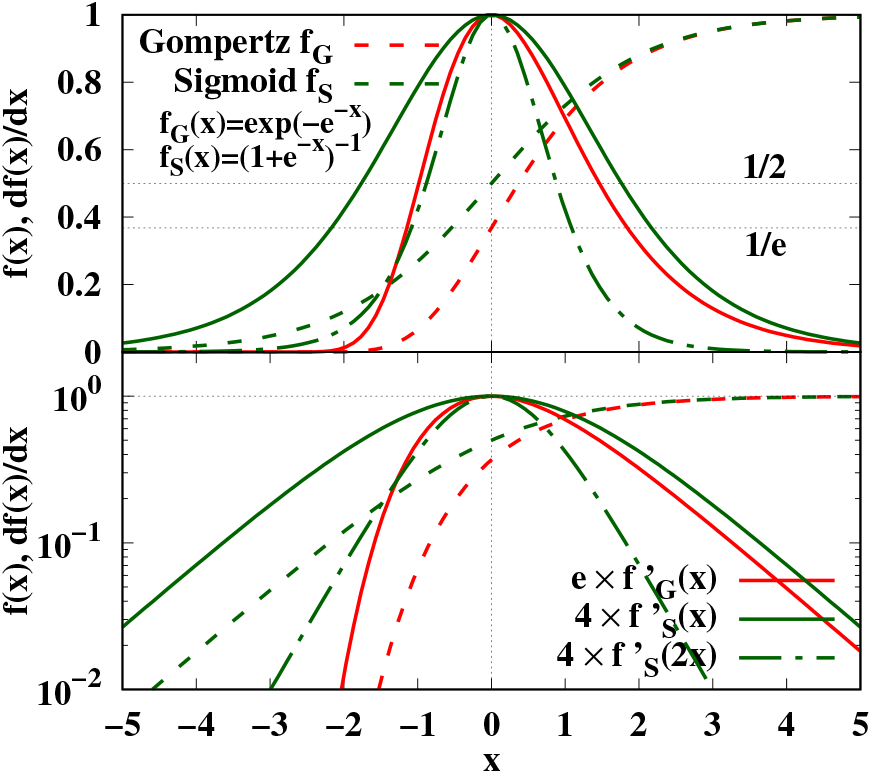
Comparison of Gompertz *f_G_*(*x*) and sigmoid *f_S_*(*x*) functions and their derivatives, 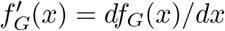 and 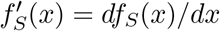, in the linear (top) and logarithmic (bottom) scales. We also show 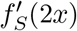, the sigmoid function with doubled infection probability, by green dot-dashed curves.

This asymmetry should be compared with the solution of the SI model [21], in which the susceptible people (*S*(*t*)) are infected by the infectious people (*I*(*t*)) at the rate proportional to the product *S*(*t*)*I*(*t*),

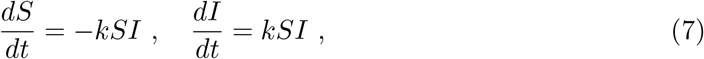

where *K* is a constant. The solution is found to be

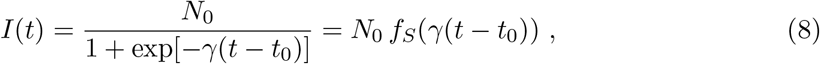

where *N*_0_ = *S* + *I* is a constant, γ = *N*_0_*k*, and *t*_0_ is the time when half of the people are infected, I(*t*_0_)= *N*_0_/2. The characteristic function *f_S_*(*x*) is called the sigmoid function, which is also referred to as the logistic function,

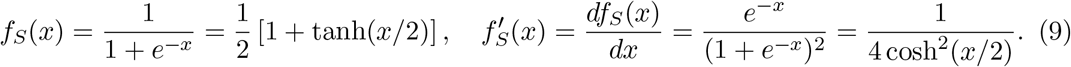

Then the daily number of infected people is a symmetric function of *t* − *t*_0_,

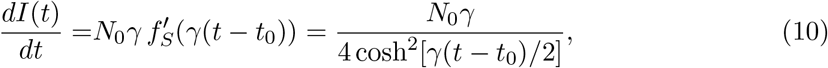

where 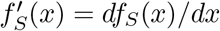. In Fig. 1, we also show *f_S_*(*x*) and *df_S_*(*x*)/*dx* by green curves. At large *x* (*x* ≥ 1), the Gompertz and sigmoid functions show similar behavior, *f_G_*(*x*) ≃ *f_S_*(*x*) ≃ 1 − exp(−*x*) and 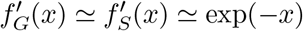. The difference found in the derivatives in the figure comes from the factor (*e* and 4) multiplied to normalize them to be unity at *x* = 0. By comparison, the sigmoid function with doubled transition probability 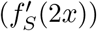) approximately agrees with 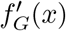 on average in the negative *x* region, −2 ≤ *x* ≤ 0. As already noticed [1–20] and is discussed later, the asymmetry is clearly found in *dN*(*t*)/*dt* for COVID-19, so *N*(*t*) is better understood by the Gompertz function than by the sigmoid function.

Once the solution is given, one can obtain the *K* value [26], increasing rate of the infected people in a week, as

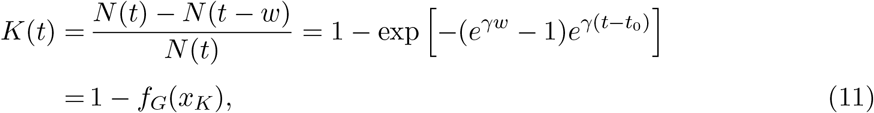

where *x_K_* is the scaling variable for *K*(*t*),

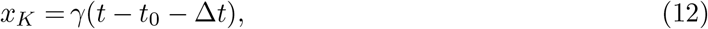

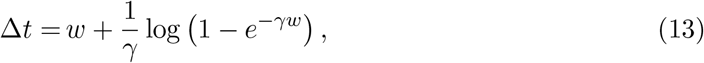

with *w* = 7 days.

In the later discussions, we regard the number of cases (the number of infected people who tested positive) as the number of infected people, since the latter is difficult to measure. We expect that the former takes a similar value to the latter, as long as the infected people are defined as the infectious people, who already have symptoms or bear enough amount of coronavirus DNAs.

## 3. Comparison with data

### 3.1. Adopted dataset

Let us now examine the universal behavior given by the Gompertz function in real data. We use the data given in Ref. [28] as of May 21, which contain the *N*(*t*) data from Dec. 31, 2019 (*t* = 0) till May 20 (*t* = 141). Throughout this article, we measure the time *t* in the unit of day. In order to avoid the discontinuity coming from the definition change, we have removed the spikes in the daily increase (*dN*(*t*)/*dt*) data in Japan (April 12, *t* = 103) and China (February 13, *t* = 44), and instead the daily numbers in previous days are increased by multiplying a common factor, which is determined to keep the total number of cases in the days after the spike. The number of cases (*N*(*t*)) is obtained as the integral of thus-smoothen *dN*(*t*)/*dt*. In addition, seven-day averages (±3 days) are considered in the analysis of *dN*(*t*)/*dt* in order to remove the fluctuations in a week. Thus-smoothen *dN*(*t*)/*dt* data are available in 3 ≤ *t* ≤ 138^1^.

We have chosen the countries with a large number of cases (the USA, Russia, Brazil, and the UK as of May 20, 2020), the first three Asian countries where the COVID-19 spread explosively (China, South Korea, and Japan), the first two countries of spread in Europe (Italy and Spain), a country with a unique policy (Sweden), and a country with somewhat different *dN*(*t*)/*dt* profile (Indonesia).

### 3.2. Number of cases and its daily increase

In the left panel of Fig. 2, we show the daily number of new cases *dN*(*t*)/*dt* given in Ref. [28] with the smoothing mentioned above. The legends stand for the abbreviation of the country name (internet country domain code, see Table 1). The *dN*(*t*)/*dt* data show there is one big peak in each country, and the shape of the peak is asymmetric; fast rise and slow decay. In many of the countries, there are several other peaks, which are smaller than the dominant one but visible at least in the log-scale plot. The fitting results using the Gompertz function are shown by dotted lines, and are found to explain the dominant peak region of data well.

**Fig. 2.**
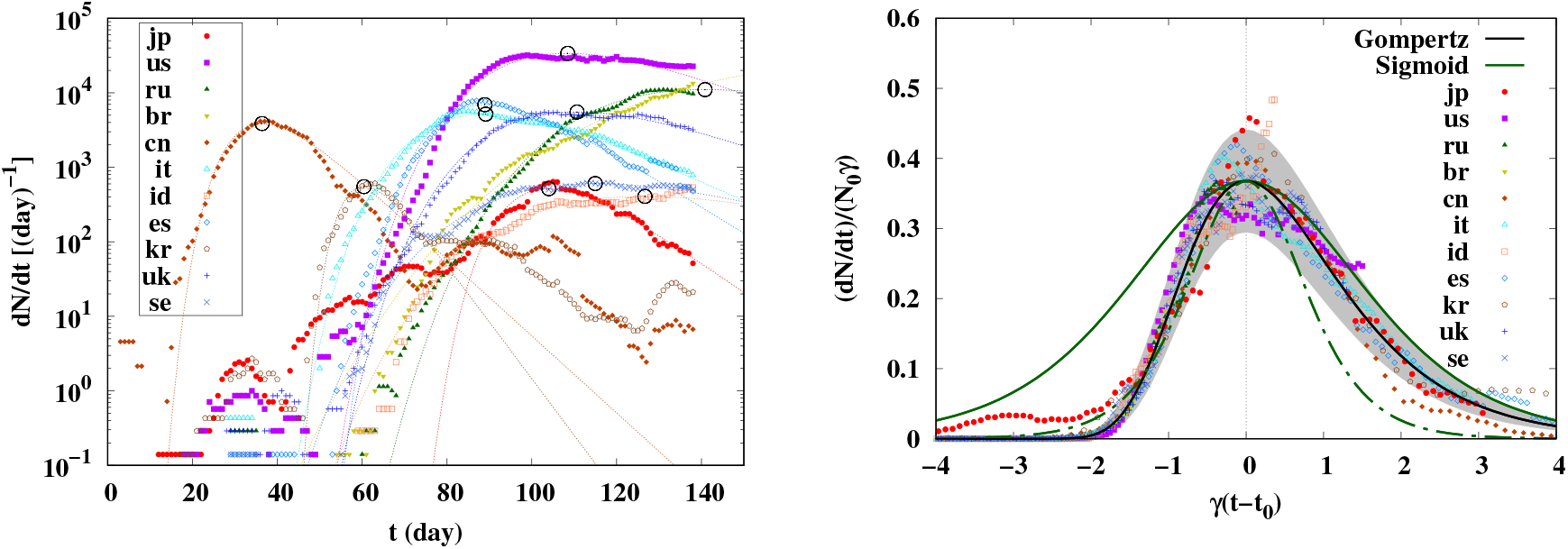
Daily number of new cases *dN*(*t*)/*dt* (left, symbols) and the scaling behavior (right, symbols). In the left panel, dotted lines show the fitting results in the derivative of the Gompertz function, 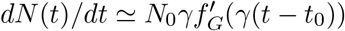, and open circles show the peak points in the fitting function, 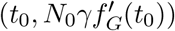. In the right panel, solid black curve shows the derivative of the Gompertz function 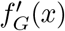, and the grey band shows its region with 5 % uncertainty in γ and 20 % in *N*_0_. The green solid (dot-dashed) curve shows the derivative of the sigmoid function normalized to reproduce the peak height, 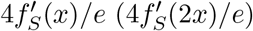.

**Table 1.** Fitting results with the fitting ranges of *t*. Parameters in the line with (*) are adopted to draw figures.

| Country | $\gamma$ [%/day] | $t_0$ [day] | $N_0$ [$10^3$] | $\Delta N$ | fit range | $\chi^2/\text{dof}$ |
| --- | --- | --- | --- | --- | --- | --- |
| Japan | $9.1 \pm 0.2$ | $104.0 \pm 0.3$ | $15.4 \pm 0.3$ | $1554 \pm 22$ | 85– 138 | 7.2 (*) |
| | $8.7 \pm 0.3$ | $103.8 \pm 0.4$ | $15.6 \pm 0.4$ | $0.5 \pm 1.3$ | 3– 138 | 11 |
| USA | $7.2 \pm 0.1$ | $100.3 \pm 0.3$ | $1202 \pm 23$ | $-800 \pm 240$ | 75– 138 | 6.7 |
| | $5.2 \pm 0.1$ | $108.5 \pm 0.4$ | $1780 \pm 28$ | $0.1 \pm 2.2$ | 3– 138 | 240 (*) |
| Russia | $3.7 \pm 0.1$ | $140.8 \pm 0.9$ | $819 \pm 28$ | $0.1 \pm 0.3$ | 3– 138 | 14 (*) |
| Brazil | $1.3 \pm 0.1$ | $245 \pm 15$ | $15100 \pm 5600$ | $-806 \pm 45$ | 74– 138 | 24 |
| | $1.9 \pm 0.1$ | $193.2 \pm 4.3$ | $3890 \pm 510$ | $-0.7 \pm 0.6$ | 3– 138 | 22 (*) |
| China | $11.9 \pm 0.2$ | $36.4 \pm 0.2$ | $88.5 \pm 1.5$ | $52.6 \pm 9.5$ | 3– 50 | 14 (*) |
| | $14.1 \pm 0.2$ | $35.4 \pm 0.2$ | $78.9 \pm 1.7$ | $73 \pm 19$ | 3– 138 | 38 |
| Italy | $6.0 \pm 0.1$ | $89.1 \pm 0.2$ | $234.6 \pm 2.0$ | $0.0 \pm 0.4$ | 3– 138 | 16 (*) |
| Indonesia | $2.7 \pm 0.2$ | $135.0 \pm 3.0$ | $42.4 \pm 3.5$ | $-173 \pm 12$ | 75– 138 | 4.6 |
| | $3.4 \pm 0.1$ | $126.7 \pm 1.2$ | $32.8 \pm 1.2$ | $-0.2 \pm 0.2$ | 3– 138 | 3.2 (*) |
| Spain | $8.1 \pm 0.1$ | $88.9 \pm 0.2$ | $231.5 \pm 2.6$ | $0.1 \pm 0.4$ | 3– 138 | 28 (*) |
| S.Korea | $17.5 \pm 0.4$ | $60.4 \pm 0.1$ | $8.5 \pm 0.2$ | $0.2 \pm 0.3$ | 3– 75 | 3.4 (*) |
| | $14.1 \pm 0.5$ | $61.1 \pm 0.3$ | $9.1 \pm 0.4$ | $0.2 \pm 0.7$ | 3– 138 | 14 |
| UK | $4.8 \pm 0.1$ | $110.6 \pm 0.4$ | $310.1 \pm 4.6$ | $20 \pm 19$ | 60– 138 | 28 |
| | $4.8 \pm 0.1$ | $110.6 \pm 0.3$ | $310.1 \pm 3.5$ | $0.1 \pm 0.4$ | 3– 138 | 16 (*) |
| Sweden | $3.5 \pm 0.1$ | $116.0 \pm 0.8$ | $46.3 \pm 1.1$ | $-36.1 \pm 5.7$ | 60– 138 | 4.3 |
| | $3.7 \pm 0.1$ | $114.9 \pm 0.6$ | $44.7 \pm 0.8$ | $-0.0 \pm 0.2$ | 3– 138 | 2.9 (*) |

The fitting to *dN*(*t*)/*dt* data is carried out by using the derivative of the Gompertz function,

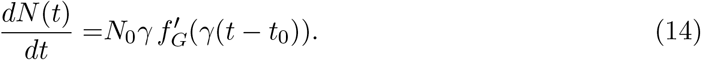

In order to concentrate on the dominant peak, we first limit the time region of the fit to *t*_min_ ≤ *t* ≤ *t*_max_, which covers it. Next, the fitting time region is extended to the whole range, 0 ≤ *t* ≤ 140. In the fitting procedure, we have assumed a Poisson distribution for the daily number of new cases, then the uncertainty in *dN*(*t*)/*dt* is assumed to be 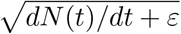, where *ε* =0.1 is introduced to avoid zero uncertainty in the case of zero daily number. We summarize the obtained parameters (*N*_0_, γ, *t*_0_) in Table 1, and parameters in the line with (*) are adopted to draw the figures. When the χ^2^ value is smaller in the whole range analysis and the obtained parameters in the two cases are similar, the single outbreak assumption is supported and we show only the results in the whole range analysis. In other cases, we in principle adopt the results giving the smaller reduced χ^2^. The exception is the USA, where the reduced χ^2^ is larger but we adopt the whole range analysis results. This is closely related to the multiple outbreaks, and will be discussed in Appendix A. It should be also noted that, unfortunately, the reduced χ^2^s are large in the single outbreak model with the present error estimate of *dN*(*t*)/*dt*. There are non-negligible contributions of other outbreaks as discussed in Appendix A. In addition, while we use the 7-day average data, we cannot completely remove the daily oscillations of *dN*(*t*)/*dt* in a week coming from the test schedule. The Gompertz function does not take care of such oscillators, then χ^2^/dof remains to be large.

In the right panel of Fig. 2, we show the normalized daily numbers, (*dN*(*t*)/*dt*)/(*N*_0_γ), as functions of the scaling variable, *x* = γ(*t* − *t*_0_). Most of the data points are around the derivative of the Gompertz function and inside the gray band, which shows the region with 5% uncertainty in γ and 20% uncertainty in *N*_0_.

We also show the sigmoid function by the green curves in the right panel of Fig. 2. It is clear that a single sigmoid function cannot describe the behavior of the *dN*(*t*)/*dt* data. If we try to fit *dN*(*t*)/*dt* data by the sigmoid function, we need to adopt larger γ in the negative *x* region as shown by the green dot-dashed curve in the right panel of Fig. 2, 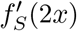, while 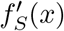 approximately agrees with 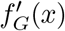 in shape in the positive *x* region.

The left panel of Fig. 3 shows the number of cases *N*(*t*), obtained as the integral of *dN*(*t*)/*dt* data after removing the spike. We also show the Gompertz function results with the parameters (*N*_0_, γ, *t*_0_) determined from the *dN*(*t*)/*dt* data, and an integration constant Δ*N*,

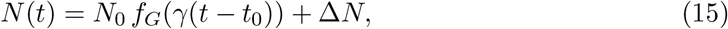

where Δ*N* is obtained by fitting to the *N*(*t*) data. In most of the countries, the Gompertz function with Δ*N* explains the data in the large *N*(*t*) region and deviations are found only in the region with small *N*(*t*). In Japan, the fitted time range is limited to be *t* ≥ 85 and earlier time data are not fitted to. Thus deviations at *t* < 80 are visible in the log scale, while the value is less than 10% of the total number of cases at *t* = 140.

**Fig. 3.**
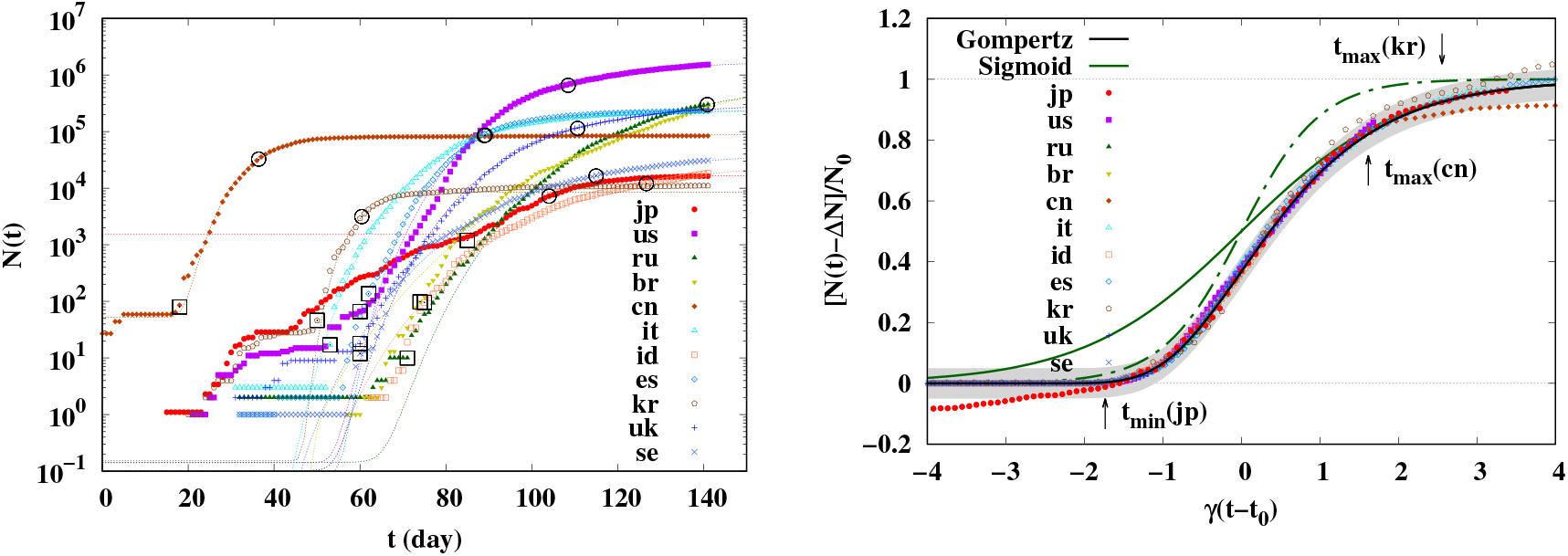
The reported number of cases *N*(*t*) (left) and the scaling behavior (right). In the left panel, dotted lines show the Gompertz function fit, *N*_0_*f_G_*(γ(*t* − *t*_0_)), open circles show the reflection points, (*t*_0_,*N*_0_/*e*), and open squares show the offset points, (*t*_offset_,*N*_offset_). In the right panel, the black solid curve shows the Gompertz function *f_G_*(*x*) and the grey band shows the region *f_G_*(*x*) ± 0.05. The green solid (dot-dashed) curve shows the sigmoid function *f_S_*(*x*)(*f_S_*(2*x*)).

We show the normalized *N*(*t*) as functions of the scaling variables in the right panel of Fig. 3. We subtract Δ*N* from *N*(*t*). It is interesting to find that most of the world data are on the Gompertz function *f_G_*(*x*). In Japan, China and South Korea, the fitted time range is limited and deviation from the Gompertz function results are found in the earlier times (Japan) and in later times (China and South Korea). It should be noted that the agreement at *x*< 0 owes largely to the large denominator compared with the number of cases in the early stage. Compared with the exponentially grown number of cases, the number of cases in the early stage is much smaller and the ratio is seen to be very small. Nevertheless, it is impressive to find the agreement of the Gompertz function and the observed number of cases after scaling.

We also show the sigmoid functions, *f_S_*(*x*) and *f_S_*(2*x*), by green solid and dot-dashed curves in the right panel of Fig. 3. As in the *dN*(*t*)/*dt* case, the sigmoid function *f_S_*(*x*) agrees with the scaled data at *x* ≥ 1, but we need to use *f_S_*(2*x*), the sigmoid function with a larger infection probability, to explain the scaled data in the region of *x* ≤ 0.

### 3.3. *K* value

We now proceed to discuss the *K* value. We choose the offset day when explosive spread started, as shown by open squares in Fig. 3. The offset day and the offset number of cases, (*t*_offset_,*N*_offset_), are summarized in Table 2. With these offset parameters, the *K* value is obtained as

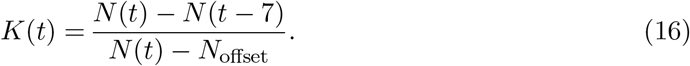

**Table 2.** Offset parameters (*t*_offset_,*N*_offset_) used to evaluate the *K* value.

| Country | $t_{\text{offset}}[\text{day}]$ | $N_{\text{offset}}$ |
| --- | --- | --- |
| Japan | 85 | 1193 |
| USA | 60 | 66 |
| Russia | 71 | 10 |
| Brazil | 74 | 98 |
| China | 18 | 80 |
| Italy | 53 | 17 |
| Indonesia | 75 | 96 |
| Spain | 62 | 136 |
| S.Korea | 50 | 46 |
| UK | 60 | 18 |
| Sweden | 60 | 12 |

Since *N*_offset_ is generally much smaller than the number of cases after explosive spread, the *K* value is not sensitive to the choice of the offset parameters as long as we discuss the long-time behavior.

In the left panel of Fig. 4, we show the *K* factor as a function of time. Data of *K*(*t*) are explained by the prediction from the scaling function, 1 − *f_G_*(*x_K_*), while the fluctuations around the predictions are large compared with the number of cases, *N*(*t*). In the right panel of Fig. 4, we show *K* values as functions of the scaling variable *x_K_* = γ(*t* − *t*_0_ − Δ*t*). Except for several countries, the scaling behavior in *K* is observed.

**Fig. 4.**
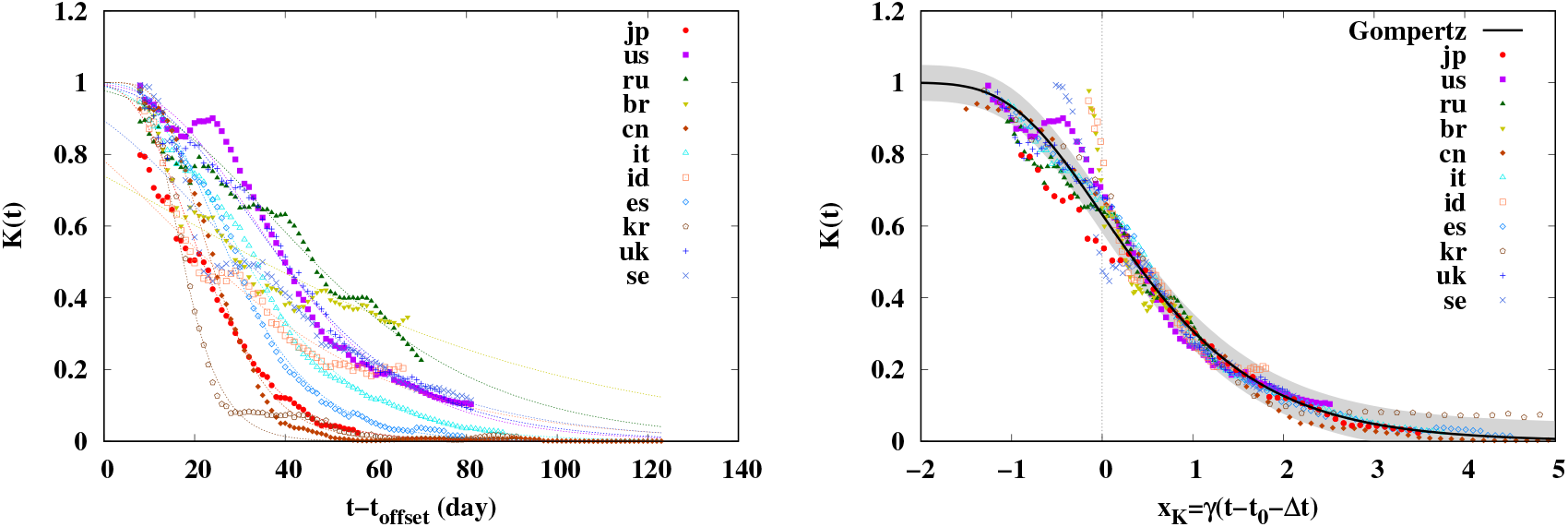
*K* value as a function of *t* (left) and as a function of the scaling variable *x_K_*. In the left panel, dotted lines show the *K* value from the Gompertz function fit. In the right panel, the black solid curve shows 1 − *f_G_*(*x_K_*) and the grey band shows the region 1 − *f_G_*(*x_K_*) ± 0.05.

In Ref. [26], *K*(*t*) is found to show the linear dependence on time in the large *K*(*t*) region, 0.25 <*K*(*t*) < 0.9. Actually, the Gompertz function *f_G_*(*x*) shows the linear dependence on time in the small |*x_K_* | region. When the scaling variable is small, the first-order Taylor expansion would work,

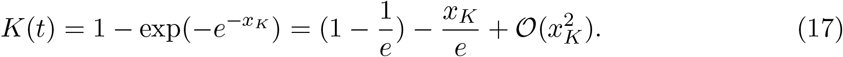

The precision of this first order approximation is around 1% (10%) for |*x_K_*|≤ 0.5(|*x_K_*|≤ 1), where 1 − *f_G_*(*x_K_*) amounts to be 0.45 − 0.81 (0.28 − 0.93). When *K* is smaller (*K*< 0.25), 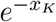 should be also small and the Taylor expansion with respect to 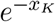 may work. Then *K* decreases exponentially,

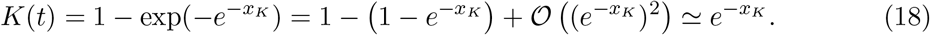

With the sigmoid function, while scaling behavior is observed in *N*(*t*) and *dN*(*t*)/*dt*, *K* does not scale as a function of a single scaling variable. In Fig. 5, we compare the functions in the *K* value derived from the Gompertz and sigmoid functions for *N*(*t*) as functions of *x* = γ(*t* − *t*_0_). With the Gompertz function, the shift of the scaling variable is enough for the *K* value to be described by the basic Gompertz function. In contrast, the *K* value from the sigmoid function reads

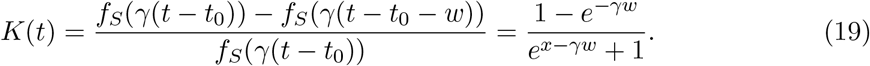

**Fig. 5.**
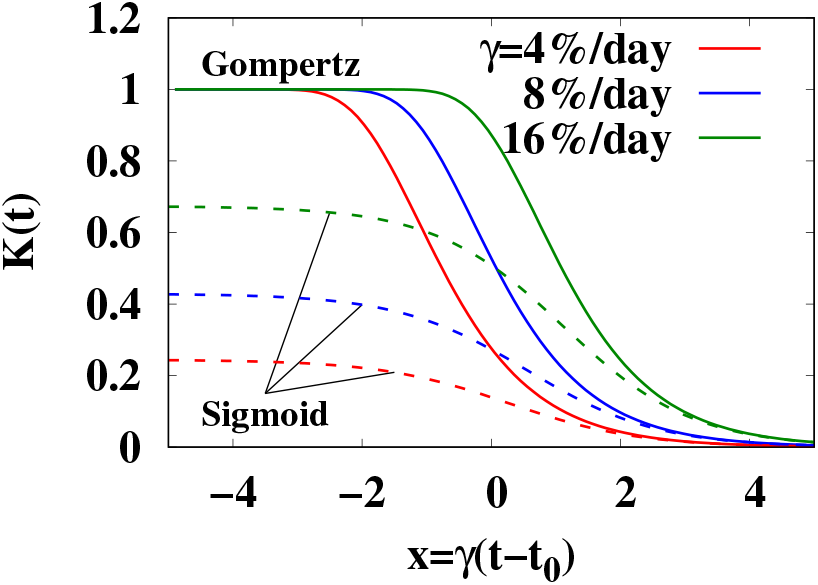
Comparison of *K* values from the Gompertz and sigmoid functions in terms of *x* = γ(*t* − *t*_0_).

In addition to the shift in the scaling variable, the amplitude also depends on γ*w* =7γ.

### 3.4. Days of expected relaxing COVID-19 restrictions from the *K* value

When the *K*(*t*) value goes down to be around 0.05 [26], it would be possible to relax COVID19 restrictions such as lifting the lockdown of the city or relaxing the state of emergency. Let us call those days as *t*_relax_. The days of relaxing the restrictions in several countries roughly correspond to the time at *K*(*t*)=0.05. With this condition, the number of cases in the current outbreak will increase by around 5% and the infection in the current outbreak will converge. In terms of the scaling variable, this corresponds to the solution of 1 − *f_G_*(*x_K_*) = 0.05 and is found to be *x_K_* = *x_R_* ≃ 2.97. Thus the expected day of relaxing the restrictions can be evaluated to be around,

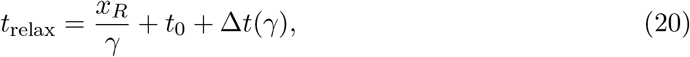

provided that there will be no further outbreaks. In Table 3, we summarize the expected day of relaxing COVID-19 restrictions *t*_relax_ from the Gompertz function analyses in comparison with the relaxed day *t*_relaxed_ in several countries.

**Table 3.** Expected day of relaxing COVID-19 restrictions *t*_relax_ and the relaxed day *t*_relaxed_.

| Country | $t_{\text{relax}}[\text{day}]$ | $t_{\text{relaxed}}[\text{day}]$ |
| --- | --- | --- |
| Japan | $135.4^{+1.5}_{-0.7}$ | 146 |
| USA | $149.9^{+2.0}_{-0.7}$ | 141 |
| Russia | $188.3^{+3.7}_{-1.2}$ | 133 |
| Brazil | $246.9^{+15.2}_{-4.2}$ | — |
| China | $63.6^{+0.8}_{-0.4}$ | 131 |
| Italy | $127.8^{+0.9}_{-0.4}$ | 125 |
| Indonesia | $175.5^{+5.2}_{-1.5}$ | — |
| Spain | $122.2^{+0.7}_{-0.3}$ | 123 |
| S.Korea | $82.4^{+0.7}_{-0.4}$ | 127 |
| UK | $153.3^{+1.5}_{-0.5}$ | 132 |
| Sweden | $162.3^{+2.7}_{-0.8}$ | — |

In Fig. 6, we show the expected days of relaxing the restrictions in comparison with some of the relaxed days. The lockdown in the Wuhan city was lifted on May 10, 2020 (*t* = 131) in China, the lockdown was relaxed on May 2, 2020 (*t* = 123) in Spain, May 4, 2020 (*t* = 125) in Italy, and May 11, 2020 (*t* = 132) in the UK. Restrictions were relaxed in part on May 6, 2020 (*t* = 127) in South Korea, May 12, 2020 (*t* = 133) in Russia, and May 20, 2020 (*t* = 141) in the USA. In Japan, the state of emergency was declared on April 7, 2020 (*t* = 98) and canceled on May 25 (*t* = 146). The relaxed days in Italy, Spain, Japan, the UK, and the USA are close to those expected from the Nakano-Ikeda model analyses. In China and Korea, the relaxed days were significantly later than the expectations from the model. One can guess that the governments tried to be on the safe side in these first two countries of COVID-19 spread.

**Fig. 6.**
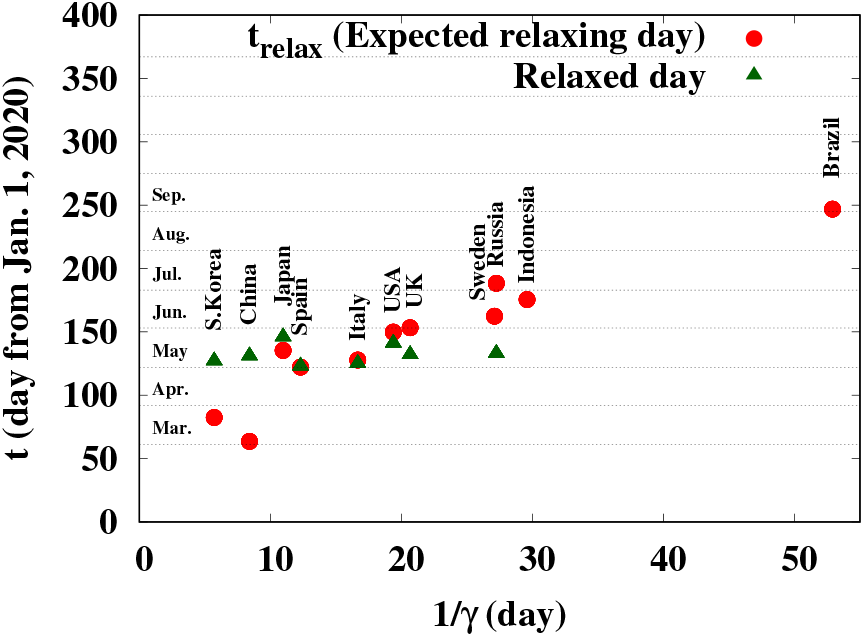
Expected days of relaxing COVID-19 restrictions and the relaxed days.

The expected day of relaxing COVID-19 restrictions strongly depends on the value of the damping rate of the infection probability, γ, which people and governments should try to enhance. In South Korea and China, the damping rate is larger than 0.1. Then the infection probability decreases by a factor of 1/*e* within 10 days. In these countries, the test-containment processes have been performed strongly. In many of the countries under consideration (Japan, USA, UK, Italy, and Spain), the damping rates take the value between 4 − 10%/day. In these countries, many of the restaurants and shops are closed and people are requested to stay home for one month or more. Sweden may be an interesting example. The Swedish government does not require restaurants and shops to be closed and does not ask people to stay home. The government asks people to be responsible for their behavior and social distancing is encouraged. The damping rate in Sweden, γ =3.7%/day, may be regarded as a value representing the intrinsic nature of COVID-19.

It would be valuable to comment on the use of the effective reproduction number, *R_e_*, which is defined as the ratio of the number of new cases in one week to that in the previous week and has the merit that it is not necessary to define the offset. For a constant infection probability per infected people, *k*(*t*)= *k*_0_ = const. in Eq. (1), *R_e_* is found to be *R_e_* = exp(*k*_0_*w*) and we can guess *k*_0_ from *R_e_*. In contrast, when the number of cases is given by the Gompertz function, *N*(*t*)= *N*_0_*f_G_*(*x*), the effective reproduction number *R_e_*(*t*) is a function of two variables, *x_K_* and γ*w*,

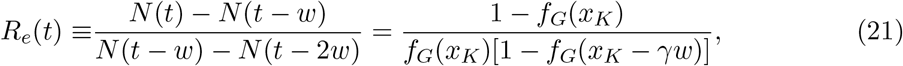

while *K*(*t*) is a function of a single scaling variable *x_K_*. Thus the universality observed in *K*(*t*) is lost in *R_e_*(*t*), and it would be less easy to give a prediction of *t*_relax_ from *R_e_*(*t*) than from *K*(*t*). In Fig. 7, we show *R_e_*(*t*) from the Gompertz function at γ =4, 8 and 16%/day. The value of *R_e_*(*t*) at *x_K_* = *x_R_* depends on γ, the value of *x_K_* at *R_e_*(*t*) = 1 is different from *x_R_*, *x_K_* =0.49 (1.42) for γ = 16% (4%)/day, and apparent large values of *R_e_*(*t*) appear in the early stage, *x_K_* < 0 or *t*<*t*_0_ +Δ*t*(γ).

**Fig. 7.**
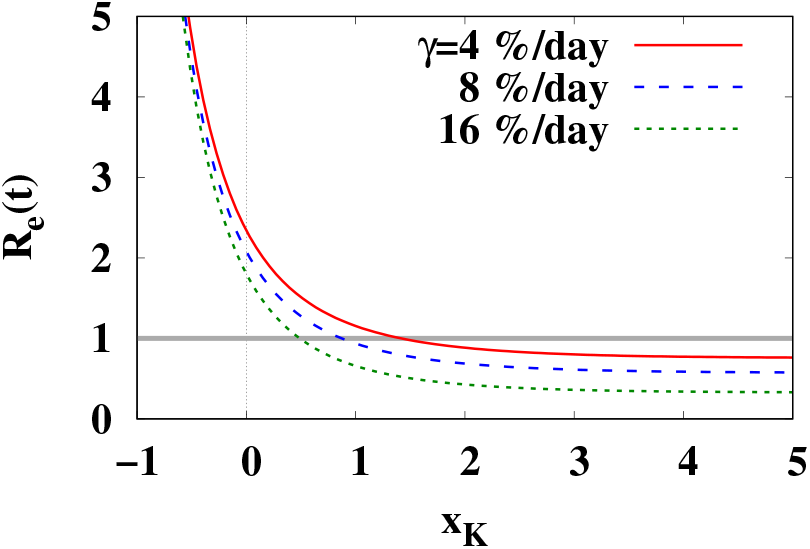
The effective reproduction number *R_e_* as a function of the scaling variable *x_K_*.

## 4. Mechanism of appearance of the Gompertz function

Fast and slow rises in the early and late stages are found in many physical processes such as the particle production in nuclear collisions. In Fig. 8, we show the number of Δ particles (Δ^++^, Δ^+^, Δ^0^ and Δ^−^) and π particles (π^+^,π^0^ and π^−^) produced in central Au+Au collisions at the incident energy of 1 GeV/nucleon [29]. Histograms show the calculated results by using the hadronic transport model JAM [30]. The main production mechanism of π particles at this incident energy is the Δ production and its decay. In the early stage (*t*< 15 fm/c), nucleons are excited to resonances such as the Δ particles in the nucleon-nucleon collisions, *NN* → *N*Δ. Produced Δs collide with other nucleons and Δs, some of them produce additional Δ particles, Δ*N* → ΔΔ, and some of them are deexcited to nucleons in the Δ absorption processes such as Δ*N* → *NN*. The Δ particles decay and produce π particles, Δ → *N*π. Produced π particles may collide with other nucleons, Δs and πs, and occasionally produce additional Δ, π*N* → πΔ. In the later stage, the system expands, particle density decreases, and interaction rate goes down. When the density becomes low enough, all Δ particles decay to *N*π with the lifetime τ_Δ_ = *ħ*/ΓΔ ≃ 2 fm/*c* and π particles go out from the reaction region and are detected.

**Fig. 8.**
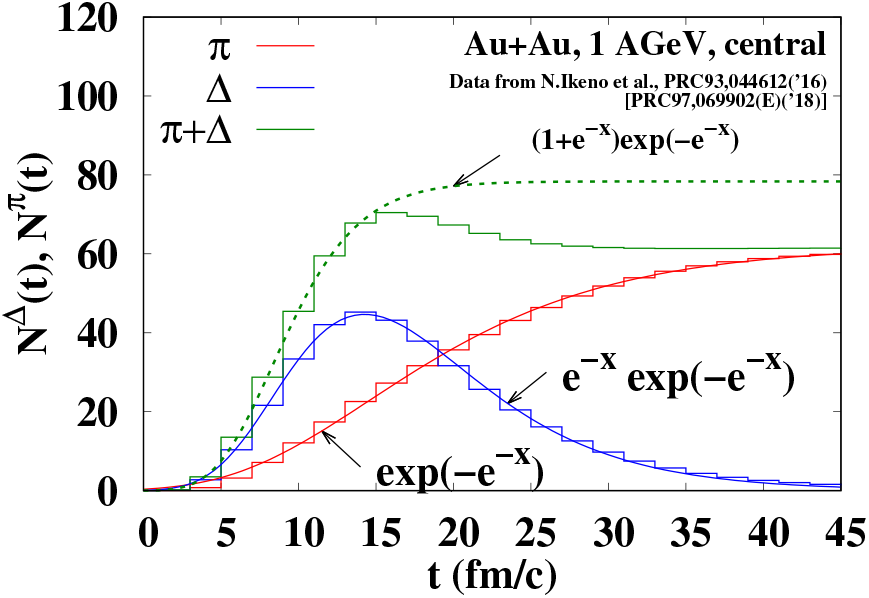
The number of π and Δ particles as functions of time *t* in central Au+Au collisions at the incident energy of 1 GeV per nucleon. Histograms show the π (red) and Δ (blue) numbers taken from Ref. [29], and solid curves show the fitting results using the Gompertz function. The green dashed curve shows the sum of π and Δ numbers.

The number of π and Δ particles in the above nucleus-nucleus collision is found to be well fitted by the Gompertz function *f_G_*(*x*) and its derivative *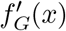*. Curves in Fig. 8 show the results of fit by the Gompertz function and its derivative,

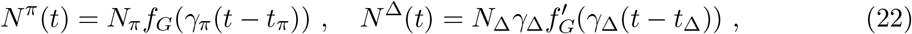

where the parameters are obtained as (*N*_π_,γ_π_,*t*_π_) = (62.0, 0.113 (fm/*c*)^−1^, 14.6 fm/*c*) and (*N*_Δ_,γ_Δ_,*t*_Δ_) = (760, 0.160 (fm/*c*)^−1^, 14.3 fm/*c*). Compared with the lifetime of Δ, the number of Δ during nucleus-nucleus collisions has a longer tail. This may be because of the relativistic effects, resonance mass dependence of the width, and sequential decay and production of Δ such as Δ → π*N* followed by π*N* → Δ.

Let us discuss the time dependence of the pion production in a simplified treatment, where the microscopic processes are limited to be the Δ production from *NN* collisions and the Δ decay, and the Markov processes are assumed,

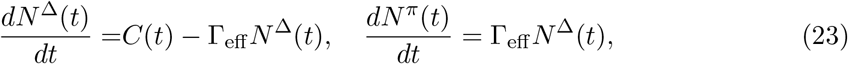

where *C*(*t*) denotes the production rate of Δ from *NN* collisions and Γ_eff_ represents the effective decay rate of Δ. The above Δ decay represents the net decay rate, the difference of the rate of Δ → *N*π and *N*π → Δ in nuclear collisions. By further assuming that Γ_eff_ is constant in time, the solution of Eq. (23) is obtained as 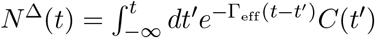. Then after the time where *C*(*t*) vanishes, *N*^Δ^ decays exponentially and its integral, *N*^π^, approaches to a constant. These features agree with those of the Gompertz function in the late stage. Earlier time behavior depends on the function form of *C*(*t*). In terms of the scaling variable, *x* =Γ_eff_ (*t* − *t*_0_), Eq. (23) reads 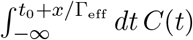. The function *N*^Δπ^ is a rapidly increasing function of time in the early stage, and the increasing rate becomes smaller in the later stage because of the energy loss and expansion of nuclear matter. If we fit the curve to the *N*^Δπ^ data in Ref. [29] by using 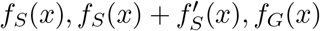 and 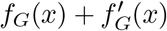 in the Δ production region (*t* ≤ 15 fm/*c*), the variance of residuals (reduced χ^2^ with the uncertainty of unity being assumed in the data) is the smallest with 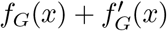 as shown by the green dashed curve in Fig. 8. This supports that *N*^π^ is described by a Gompertz function, while the mechanism of this function form to appear is not understood. It should be noted that the number of Δ is proportional to the increasing rate of π number in the present simplified treatment, but the fitting to the transport model data results in γ_Δ_ ≠ γ_π_, implying that this is not the case.

It should be noted that one Δ particle mostly decays into *N*π and additionally produced number of π is less than unity in the present pion production in nucleus-nucleus collisions. A rough estimate of the upper bound of the basic reproduction number *R*_0_ from Δ to π may be obtained as follows. The lower bound of the number of produced Δ particles is the peak number of Δ, which is *N*^Δ^ = 45.3 at *t* = 14 fm/*c* in the calculated data and is expected to be *N*^Δ^(*t*_Δ_)= *N*_ΔγΔ_/*e* ≃ 44.7 at *t* = *t*_Δ_ from the Gompertz function. Then the upper bound of the additionally produced π number is given as *N*^π^(*t* = ∞) − *N*^Δ^(*t*_Δ_) ≃ 17. Consequently, the upper bound of *R*_0_ is given as *R*_0_ =[*N*^π^(*t* = ∞) − *N*^Δ^(*t*_Δ_)]/*N*^Δ^(*t*_Δ_) ≃ 0.38, which is less than unity. As a result, the number of pions in the final state is already determined in the early stage. The green dashed curve in Fig. 8 shows the “π-like” particle number, *N*^πΔ^ = *N*^π^ + *N*^Δ^, which shows a peak at *t* ≃ 16 fm/*c*, gradually decreases by the Δ absorption processes, and converges to *N*^π^(*t* = ∞).

Based on the success in describing *N*^π^(*t*) by using the Gompertz function as in the case of *N*(*t*) associated with COVID-19, we may make a conjecture of the correspondence of the COVID-19 spread and the π production in nuclear collisions. Let us assume that π particles correspond to the cases. Then Δ particles are regarded as the coronavirus carriers, defined as the infected people who have not tested positive yet. Carriers will test positive later or will be recovered without testing positive. The susceptible people are infected and become carriers in the initial dense stage (Δ production), occasionally infect other susceptible people (additional Δ production) or are recovered (Δ absorption), develop symptoms and test positive (Δ → *N*π). Thus the number of carriers (*N*^Δ^) increases rapidly by explosive spread of infection in the early stage (Δ production stage), while it decreases more slowly by developing symptoms in the late stage (Δ decay stage). This causes the asymmetric time-profile in the number of carriers (*N*^Δ^), which is roughly proportional to the daily number of new cases (*dN*^π^/*dt*). By comparison, the number of infected people including carriers (*N*^π^ + *N*^Δ^) grows rapidly in the early dense stage but does not change much in the later stage. Hence, except for the early dense stage, the basic reproduction number would be less than unity.

## 5. Summary

We have analyzed the number of cases (the number of infected people who tested positive for COVID-19), as a function of time, *N*(*t*), by using the double exponential function referred to as the Gompertz function, *f_G_*(*x*) = exp(−*e*^−^*^x^*). The Gompertz function appears when the infection probability is an exponentially decreasing function of time. One of the characteristic features of the Gompertz function is the asymmetry of its derivative, 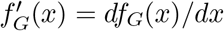, fast rise and slow decay.

This feature is found in the daily new cases, *dN*(*t*)/*dt*. We have assumed that the number of cases from one outbreak is given as 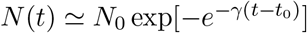, where *N*_0_, γ and *t*_0_ are the final number of cases, damping rate of the infection probability, and the time where the daily number of new cases peaks out. These parameters are obtained by the χ^2^ fitting to the *dN*(*t*)/*dt* data. Then we have found that *N*(*t*) and *dN*(*t*)/*dt* show universal scaling, *N*(*t*)/*N*_0_ = *f_G_*(*x*) and 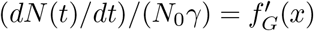, where *x* = γ(*t* − *t*_0_) is the scaling variable. The *K* value, the increasing rate of cases in one week, is also found to show the scaling behavior, *K*(*t*)=1 − *f_G_*(*x_K_*), where *x_K_* = γ(*t* − *t*_0_ − Δ*t*(γ)) is the scaling variable for *K*(*t*) with Δ*t*(γ) being a given function of γ.

We have also found that the time dependence of the produced pion number in nucleus-nucleus collisions is described by the Gompertz function. Since both of the COVID-19 spread and the pion production are transport phenomena, the mechanism of the former may be similar to the latter. If this is the case, there is a possibility that the basic reproduction number is high only in the initial stage of the outbreak.

Throughout this article, we have imposed the single-outbreak assumption. Since this assumption may be too restrictive, we show the results of multiple-outbreak analyses in Appendix A. The multiple-outbreak analyses also show that the COVID-19 spread in one outbreak is well described by the Gompertz function. We also note that after submitting the original manuscript of this article, the daily numbers of new cases are found to be significantly larger than the predictions given in Fig. 2 (and Fig. A1 in Appendix A). We give brief descriptions of the data observed later in some countries in Appendix B.

## Data Availability

We use data opened to public and available on the web.
https://ourworldindata.org/coronavirus-source-data
https://covidtracking.com/

## Acknowledgments

The authors thank T. Kunihiro and T. T. Takahashi for constructive comments and suggestions. They also thank N. Ikeno, A. Ono and Y. Nara for useful discussions. This work is supported in part by the Grants-in-Aid for Scientific Research from JSPS (Nos. 19H01898 and 19H05151).

## A. Multiple-outbreak model analysis

In the analyses in the main text, we have assumed that there is only one dominant outbreak. Let us consider here the multiple-outbreak cases, where we assume the number of cases is described by the sum of several Gompertz functions,

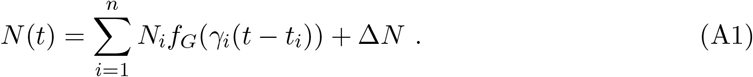

As in the single-outbreak model discussed in the main text, we analyze the daily number of new cases, *dN*(*t*)/*dt*,

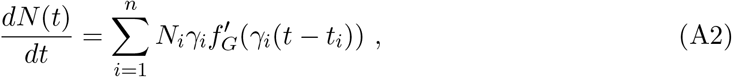

where 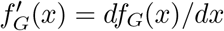.

In Fig. A1, we show *dN*/*dt* in Eq. (A2) in comparison with the daily number of new cases. In the multiple-outbreak analysis, we use data in the whole range, 0 ≤ *t* ≤ 140. Two or three outbreaks are considered, and we try to describe the region with large *dN*/*dt* by adding outbreaks. Obtained parameters are summarized in Table A1.

In many countries under consideration, *dN*/*dt* is decreasing on May 21, 2020, and the parameters are well determined. Then we adopt the three-outbreak model (*n* = 3). In Japan, China, and South Korea, multiple-outbreak structure of *dN*/*dt* is clearly seen in the logarithmic plot and can be fitted by using Eq. (A2). In Russia, Italy, Spain, the UK and Sweden, additional outbreaks improve the reduced χ^2^ by filling the peaks which are not covered by the single outbreak. In Brazil and Indonesia, where the numbers of cases are still rapidly increasing, we need at least one outbreak term with *t_i_* >*t*_now_. In those cases, parameters generally have large uncertainties, so the third outbreak, if included, has extremely large uncertainties larger than 100 %. Thus we use the two-outbreak model (*n* = 2) in these countries.

In the USA, there are many centers of outbreaks. In Fig. A2, we show the *dN*(*t*)/*dt* data in New York, Massachusetts and California states [31]. It is possible to fit the data in each state by using the Gompertz function, but the results show significantly different values in (γ, *t*_0_). This would be the reason of the slow decrease of *dN*(*t*)/*dt* in the USA. As a result, a single-outbreak treatment is not appropriate. By comparison, the *dN*(*t*)/*dt* data are reasonably explained by two outbreaks (*n* = 2). We have used the data in Ref. [28] updated on June 7, 2020. The daily number of new cases does not decrease and it seems that it takes more time for the settle down.

**Fig. A1.**
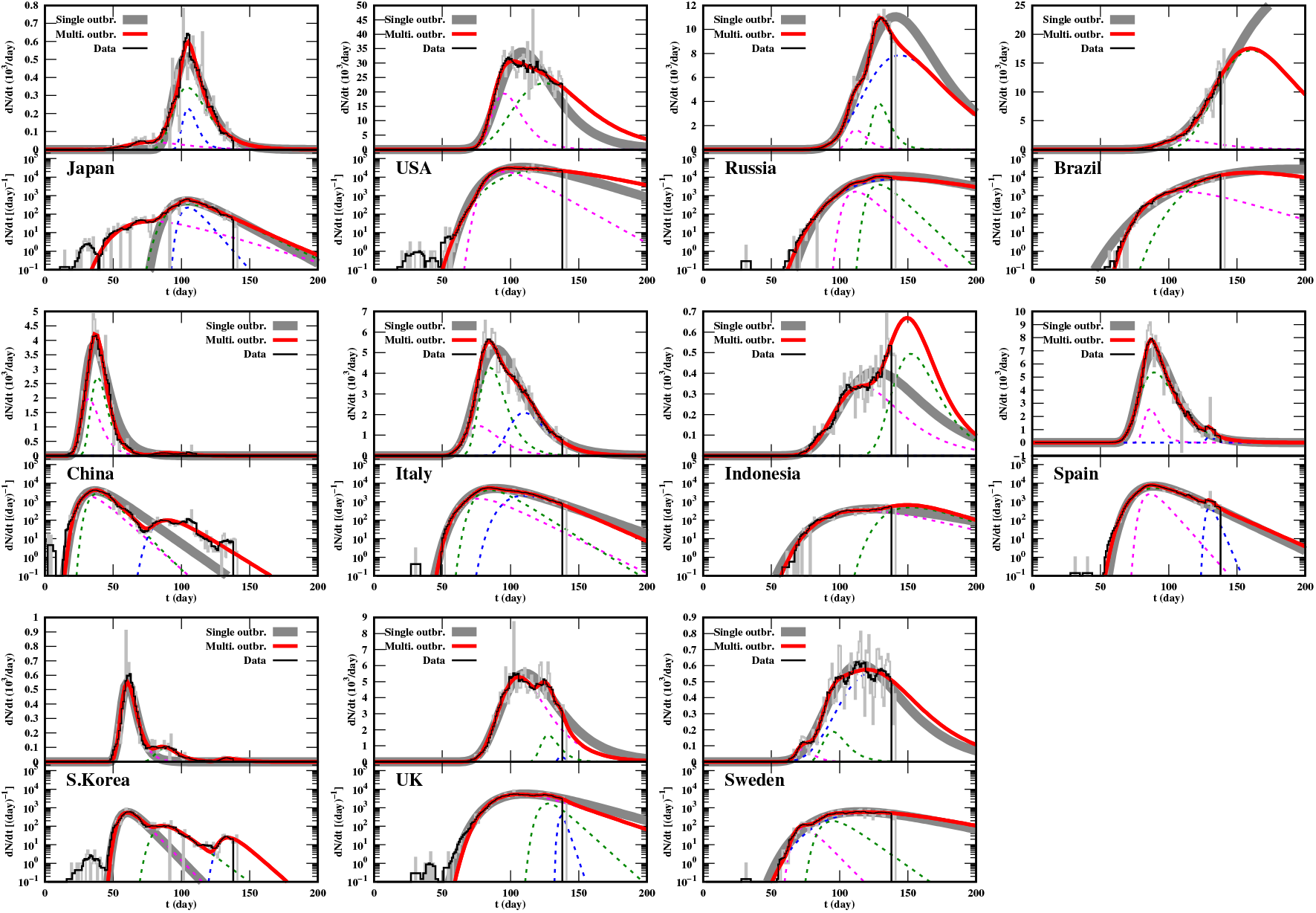
Daily number of new cases *dN*/*dt* with single-outbreak (grey) and multiple-outbreak (red) model analyses in the linear (top) and logarithmic (bottom) scales. Magenta, green and blue dotted curves show the contributions from the first, second and third outbreaks, respectively.

**Fig. A2.**
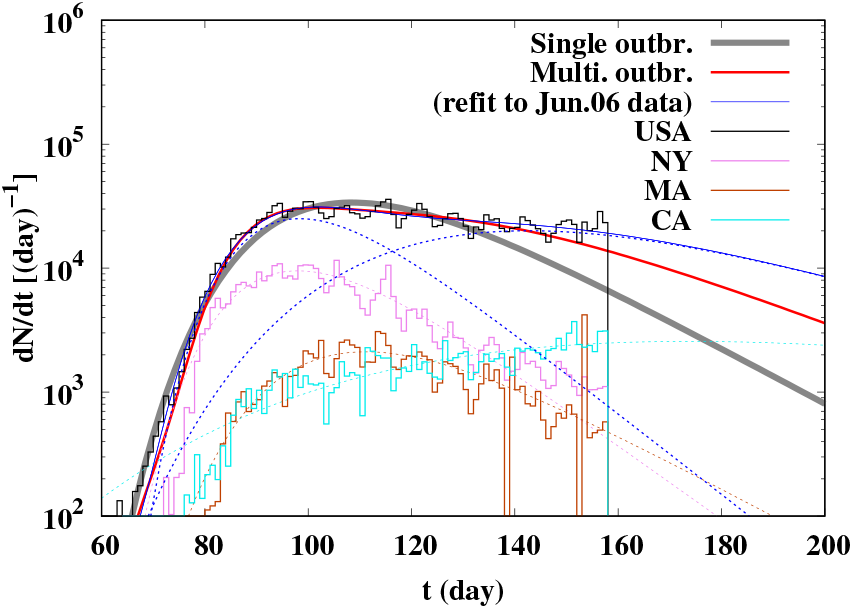
Daily number of new cases *dN*(*t*)/*dt* in the USA with single-outbreak (grey) and multiple-outbreak (red and blue) model analyses. The red and blue curves show the fitting results to the data till May 20 and June 6, respectively. Magenta, brown and cyan histograms (curves) show the data (fitting results in the single-outbreak model) in the New York, Massachusetts, and California states, respectively.

**Table A1.** Parameters in multiple-outbreak model analyses.

| Country | $\gamma_i$[%/day] | $t_i$[day] | $N_i$[$10^3$] | $\Delta N$ |
| --- | --- | --- | --- | --- |
| Japan | $5.0 \pm 0.5$ | $78.5 \pm 3.0$ | $2.2 \pm 0.3$ | $0.3 \pm 0.3$ |
| | $8.2 \pm 0.3$ | $104.3 \pm 0.2$ | $11.4 \pm 0.6$ | |
| | $19.4 \pm 1.8$ | $105.0 \pm 0.3$ | $3.2 \pm 0.6$ | |
| USA | $9.4 \pm 0.2$ | $95.4 \pm 0.4$ | $560 \pm 33$ | $0.2 \pm 0.5$ |
| | $3.7 \pm 0.1$ | $124.8 \pm 1.2$ | $1682 \pm 21$ | |
| Russia | $15.4 \pm 0.5$ | $111.8 \pm 0.2$ | $28.9 \pm 1.8$ | $0.1 \pm 0.1$ |
| | $16.1 \pm 0.6$ | $128.8 \pm 0.2$ | $65.8 \pm 5.1$ | |
| | $3.3 \pm 0.0$ | $144.1 \pm 0.9$ | $647 \pm 31$ | |
| Brazil | $5.0 \pm 0.4$ | $111.3 \pm 3.5$ | $86.6 \pm 22.4$ | $0.0 \pm 0.1$ |
| | $3.4 \pm 0.4$ | $160.6 \pm 4.9$ | $1380 \pm 330$ | |
| China | $14.8 \pm 2.1$ | $32.0 \pm 1.9$ | $36.5 \pm 14.6$ | $52.2 \pm 6.6$ |
| | $17.0 \pm 1.3$ | $38.5 \pm 0.3$ | $43.8 \pm 14.6$ | |
| | $10.4 \pm 0.6$ | $90.0 \pm 0.8$ | $2.5 \pm 0.2$ | |
| Italy | $8.3 \pm 0.5$ | $76.6 \pm 1.8$ | $47.3 \pm 11.1$ | $0.0 \pm 0.1$ |
| | $10.6 \pm 0.4$ | $85.0 \pm 0.3$ | $110 \pm 12$ | |
| | $7.4 \pm 0.1$ | $110.0 \pm 0.6$ | $76.7 \pm 3.7$ | |
| Indonesia | $4.1 \pm 0.1$ | $116.5 \pm 1.0$ | $22.3 \pm 1.0$ | $-0.1 \pm 0.1$ |
| | $6.0 \pm 2.4$ | $152 \pm 12$ | $23 \pm 23$ | |
| Spain | $19.1 \pm 0.9$ | $86.3 \pm 0.2$ | $36.9 \pm 3.2$ | $0.1 \pm 0.2$ |
| | $7.5 \pm 0.1$ | $89.5 \pm 0.1$ | $195.4 \pm 3.2$ | |
| | $39.8 \pm 3.1$ | $129.9 \pm 0.2$ | $3.6 \pm 0.3$ | |
| S.Korea | $17.5 \pm 0.3$ | $60.4 \pm 0.1$ | $8.5 \pm 0.2$ | $0.2 \pm 0.2$ |
| | $12.6 \pm 0.6$ | $87.8 \pm 0.5$ | $2.0 \pm 0.1$ | |
| | $15.0 \pm 3.7$ | $134.1 \pm 1.9$ | $0.4 \pm 0.1$ | |
| UK | $5.7 \pm 0.1$ | $106.1 \pm 0.3$ | $251.5 \pm 3.5$ | $0.1 \pm 0.3$ |
| | $15.5 \pm 1.3$ | $127.6 \pm 0.4$ | $29.4 \pm 3.0$ | |
| | $50 \pm 38$ | $137.0 \pm 1.4$ | $2.0 \pm 1.9$ | |
| Sweden | $17.2 \pm 1.4$ | $73.2 \pm 0.5$ | $1.3 \pm 0.2$ | $0.0 \pm 0.1$ |
| | $12.0 \pm 1.1$ | $94.9 \pm 0.7$ | $4.3 \pm 0.8$ | |
| | $3.4 \pm 0.1$ | $124.1 \pm 1.7$ | $44.6 \pm 0.9$ | |

After obtaining (*N_i_*,γ*_i_*,*t_i_*) by fitting *dN*/*dt* data, the constant part (Δ*N*) is obtained by fitting *N*(*t*). Thus obtained multiple-outbreak functions in Eq. (A1) are compared with the data in Fig. A3. In the region with *N*(*t*) > 100, the multiple-outbreak functions are found to explain the data well. This supports the idea that the number of cases in one outbreak would be described by the Gompertz function.

**Fig. A3.**
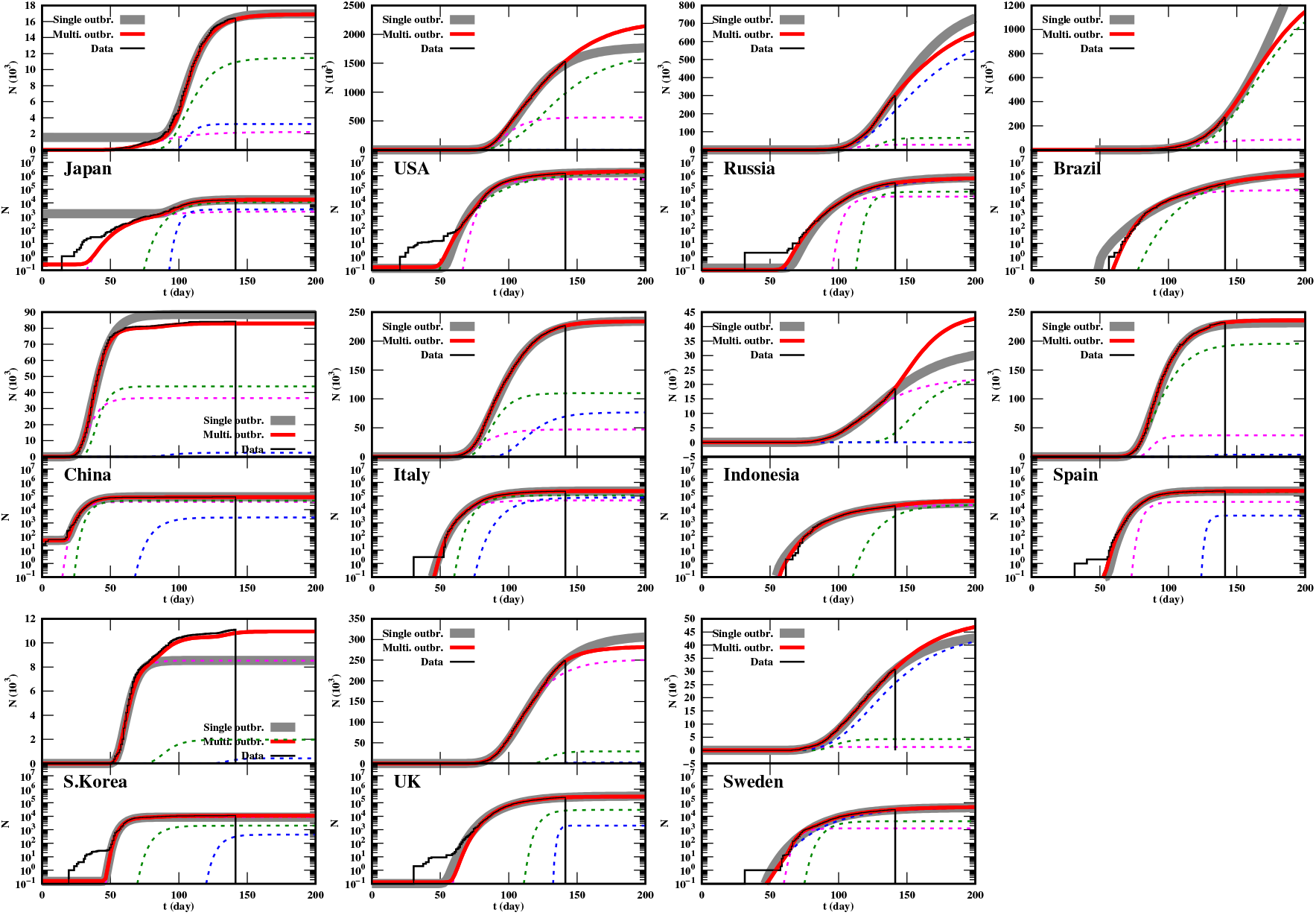
Number of cases *N*(*t*) with single-outbreak and multiple-outbreak model analyses in the linear (top) and logarithmic (bottom) scales. Magenta, green and blue dotted curves show the contributions from the first, second and third outbreaks, respectively.

## B. COVID-19 spread in July and August

After submitting the original manuscript in June, 2020, we find excess of cases over the predictions in several countries. It is also valuable to verify the consequences of the lockdowns. We give a brief description of the above mentioned issues. This Appendix is given as a “note added”, and the details will be reported elsewhere.

In Sweden, the USA and Japan, significant excess of cases over the Gompertz function analyses has been observed after mid June as shown in Fig. B1. In addition to *dN*(*t*)/*dt*, *dD*(*t*)/*dt* can be also explained by the Gompertz functions [1], as shown by blue (*dN*(*t*)/*dt*) and red (*dD*(*t*)/*dt*) solid curves. Each of the dotted curves shows the contribution from one outbreak. We have used two and three Gompertz function terms for Sweden and the USA, respectively, and three and four terms for *dN*(*t*)/*dt* and *dD*(*t*)/*dt* in Japan. In Sweden, *dN*(*t*)/*dt* started to increase again around *t* = 140, but *dD*(*t*)/*dt* kept decreasing. In the USA, *dN*(*t*)/*dt* started to increase again around *t* = 165, Compared with the new cases, the increase of *dD*(*t*)/*dt* after mid June is less prominent. The grey dashed line shows the fitting results of the new cases shifted later by seven days and multiplied by 0.07, *A* × *dN*(*t* − *t_D_*)/*dt* with *A* =0.07 and *t_D_* = 7. This curve roughly agrees with *dD*(*t*)/*dt* until *t* ≃ 130 but is larger than *dD*(*t*)/*dt* later. A similar trend is observed but more strongly in Japan. We find *dN*(*t*)/*dt* once decreased rapidly but increased again around *t* = 160. The shifted and scaled function of the new cases (*A* =0.05 and *t_D_* = 14) roughly explains the daily number of deaths until *t* ≃ 170 but it overestimates *dD*(*t*)/*dt* later.

**Fig. B1.**
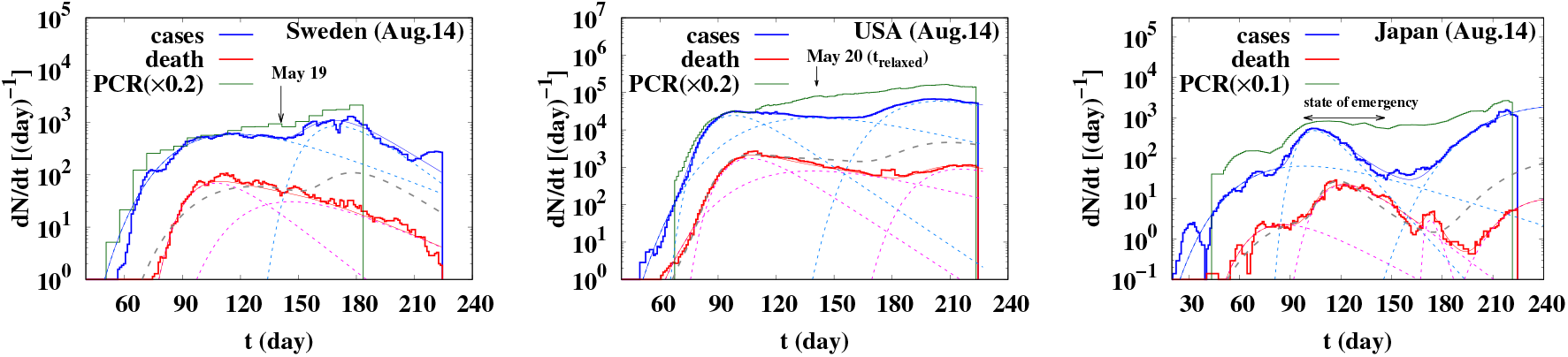
The number of new cases *dN*(*t*)/*dt* (blue histograms), the daily number of deaths *dD*(*t*)/*dt* (red histograms), and the number of new PCR tests with the rescaling factor 0.1 or 0.2 (green histograms) in Sweden (left), the USA (middle) and Japan (right). Histograms show the data taken from Ref. [28]. Solid (dotted) curves show the fitting results in the multiple-outbreak model for *dN*(*t*)/*dt* (blue) and *dD*(*t*)/*dt* (red) (their breakdowns). The grey dashed curves show the shifted and scaled *dN*(*t*)/*dt* results in the model (see the text).

Let us guess the reason of these behaviors. There are several possible reasons of the *dN*(*t*)/*dt* increase in July, such as (a) increase of the number of PCR tests and improved sensitivity, (b) lifting the lockdown and economic resumption, and (c) genomic mutation [32]. As for the decrease of mortality (death rate), (ab’) dominance by young and mild symptom cases from (a) and (b) with improved medical care, (c) genomic mutation [32], and (d) *t* cell immunity [33] are the candidate reasons. In Sweden, the government expanded coronavirus testing to include people with mild symptoms on May 19 (*t* = 141), then the increase of cases after *t* = 140 may have been caused mainly by the candidate reason (a), the increase of the number of PCR tests. The increase of cases would have been dominated by the people with mild symptoms and the actual number of infected people may have not increased. Then it is reasonable that *dD*(*t*)/*dt* did not increase. In the USA and Japan, *dD*(*t*)/*dt* also started to increase in late June (*t* ≥ 160), and then the reason (a) is not enough. We guess that the candidate reason (b), lifting the lockdown and economic resumption, would have affected the spread, and that (ab’), dominance by young and mild symptom cases with improved medical care, may have suppressed the mortality. We would like to mention other possibilities, (c) and (d). It is already pointed out that the genomes with advanced mutation are found after late June in Japan [32]. The candidate reason (c), a genomic mutation, may have caused the July and August epidemic and reduced mortality. Another candidate is (d), the *t* cell immunity [33]. The memory *t* cells exposed by the SARS-CoV-2 may work for long-term immune protection against COVID-19 [33], and these effects may be enhanced by the BCG vaccination [34]. This mechanism can be universal and explains the suppression of mortality in many countries. The actual mechanism of the present coronavirus spread would be the combination of (a)-(d) and others. Analyses of more data and further research of infection diseases are necessary to elucidate the mechanism. These are out of the scope of this article.

**Fig. B2.**
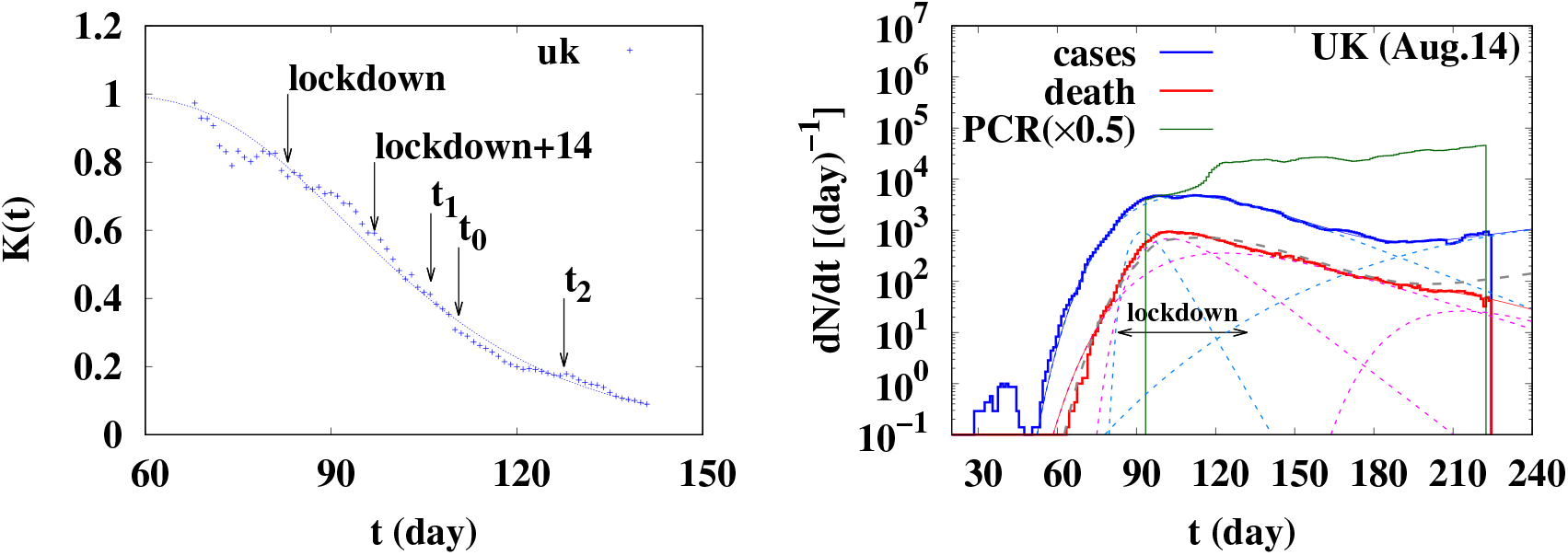
The *K* value as a function of *t* (left) and the number of new cases *dN*(*t*)/*dt*, the daily number of deaths *dD*(*t*)/*dt*, and the number of new PCR tests with the rescaling factor 0.5 (right) in the UK. The meaning of the curves in the right panel are the same as those in Fig. B1. In the left panel, we show the lockdown time, two weeks later after the lockdown, the peak time(s) in the single (multiple) outbreak model *t*_0_ (*t*_1_ and *t*_2_) by arrows. The grey dashed line in the right panel shows the shifted and scaled function of the new cases with *A* =0.15 and *t_D_* = 7.

The numbers of total confirmed cases and the *K* value should depend on both the lockdown of the social systems and the number of PCR testing carried out in the countries. Let us here take a look at these effects in the case of the UK, as an example. In Fig. B2, we show the *K* value before *t* = 140 (left) and the number of new cases, death and PCR tests (right). The peak time was *t* = *t*_0_ = 110.6(*t* = *t*_1_ = 106.1) in the single-outbreak (multiple-outbreak) analysis. The lockdown in the UK started on March 23 (*t* = 83), and the *K* value started to decrease around two weeks after the lockdown (*t* = 97) as seen in the left panel of Fig. B2, while the decrease is not enough to go well below the Gompertz curve. Nevertheless, we should mention that the lockdown prevented the next outbreak to start. The increase of new cases around *t* = 120 is considered to be caused by the speedy increase of the PCR testing since the daily number of deaths kept decreasing, and the next outbreak started later than the lift of the lockdown, *t* = *t*_relaxed_ = 132. It should be noted that these are conjectures deduced from the *dN*(*t*)/*dt* and *dD*(*t*)/*dt* data, and confirmation by further analyses would be necessary.

## Footnotes

1 In actual fitting processes, we regard the daily difference as the derivative, *N*(*t*) − *N*(*t* − 1) = *dN*/*dt*|*_t_*_−1/2_, then the actual fitting range is 2.5 ≤ *t* ≤ 137.5 for *dN*/*dt*. In the following discussions, this half-day difference is not explicitly written but should be understood.

## References

[1] M. Levitt, A. Scaiewicz, F. Zonta, medRxiv (2020), https://doi.org/10.1101/2020.06.26.20140814; Michael Levitt, online lectures on May 15th, 2020, linked from http://med.stanford.edu/levitt.html (2020).

[2] P. Castorina, A. Iorio, and D. Lanteri (2020), arXiv:2003.00507 [physics.soc-ph].

[3] Lin Jia, Kewen Li, Yu Jiang, Xin Guo, and Ting Zhao (2020), arXiv:2003.05447 [q-bio.PE].

[4] Giuseppe Dattoli, Emanuele Di Palma, Silvia Licciardi, and Elio Sabia (2020), arXiv:2003.08684 [q-bio.PE].

[5] D. Lanteri, D. Carco’, and P. Castorina (2020), arXiv:2003.12457 [q-bio.PE].

[6] José Miguel Ponciano, Juan Adolfo Ponciano, Juan Pablo Gómez, Robert D. Holt, and Jason K. Blackburn (2020), arXiv:2005.11201 [q-bio.PE].

[7] Jiri Mazurek and Zuzana Nenickova (2020), https://doi.org/10.13140/RG.2.2.19841.81761.

[8] Wuyue Yang, Dongyan Zhang, Liangrong Peng, Changjing Zhuge, and Liu Hong, medRxiv (2020), https://doi.org/10.1101/2020.03.12.20034595.

[9] Ali Ahmadi, Yasin Fadaei, Majid Shirani, and Fereydoon Rahmani, medRxiv (2020), https://doi.org/10.1101/2020.03.17.20037671.

[10] Yuri Tani Utsunomiya, Adam Taiti Harth Utsunomiya, Rafaela Beatriz Pintor Torrecilha, Silvana Cassia Paulan, Marco Milanesi, and Jose Fernando Garcia, medRxiv (2020), https://doi.org/10.1101/2020.03.30.20047688.

[11] Daren J Austin, medRxiv (2020), https://doi.org/10.1101/2020.04.09.20059402.

[12] Jean Roch Donsimoni, René Glawion, Bodo Plachter, Constantin Weiser, and Klaus Wälde, medRxiv (2020), https://doi.org/10.1101/2020.04.10.20060301.

[13] Thomas Klabunde and Clemens Giegerich, medRxiv (2020), https://doi.org/10.1101/2020.04.14.20064790.

[14] Renato Rodrigues Silva, Wisley Donizetti Velasco, Wanderson da Silva Marques, and Carlos Augusto Goncalves Tibirica, medRxiv (2020), https://doi.org/10.1101/2020.04.19.20071852.

[15] Carlos Maximiliano Dutra, medRxiv (2020), https://doi.org/10.1101/2020.04.22.20074898.

[16] A M C H Attanayake, sanjeewa Perera, and Saroj Jayasinghe, medRxiv (2020), https://doi.org/10.1101/2020.05.04.20091132.

[17] Marti Catala, Sergio Alonso, Enrique Alvarez Lacalle, Daniel Lopez, Pere-Joan Cardona, and Clara Prats, medRxiv (2020), https://doi.org/10.1101/2020.05.13.20101329.

[18] Rhodri P Hughes and Dyfrig A Hughes, medRxiv (2020), https://doi.org/10.1101/2020.05.15.20102764.

[19] Christophe Z Z Guilmoto, medRxiv (2020), https://doi.org/10.1101/2020.05.17.20097410.

[20] Nicola Bartolomeo, Paolo Trerotoli, and Gabriella Serio, medRxiv (2020), https://doi.org/10.1101/2020.05.20.20108241.

[21] D Dagon, C Zou, W Lee, Proc. 13th Annual Network and Distributed System Security Symposium (NDSS’06), Feb., pp 235-249, 2006.

[22] B. Gompertz, Philosophical Transactions of the Royal Society of London 115, 513 (1825).

[23] A. K. Laird, British Journal of Cancer 13, 490 (1964).

[24] K. Ohishi, H. Okamura, and T. Dohi, Journal of Systems and Software 82, 535 (2009).

[25] R. Chawla and M. Kaur, Adv. High Energy Phys. 2018, 5129341 (2018).

[26] T. Nakano and Y. Ikeda, medRxiv (2020), https://doi.org/10.1101/2020.04.25.20080200.

[27] Y. Akiyama (2020), in the note linked from http://www.bi.cs.titech.ac.jp/COVID-19/.

[28] Coronavirus Source Data, https://ourworldindata.org/coronavirus-source-data (2020).

[29] N. Ikeno, A. Ono, Y. Nara, and A. Ohnishi, Phys. Rev. C 93, 044612 (2016) [Erratum: Phys. Rev. C, 97, 069902(E) (2018)].

[30] Y. Nara, N. Otuka, A. Ohnishi, K. Niita, and S. Chiba, Phys. Rev. C 61, 024901 (2000).

[31] The COVID Tracking Project, https://covidtracking.com/ (2020).

[32] Tsuyoshi Sekizuka et al., medRxiv (2020), https://www.medrxiv.org/content/10.1101/2020.07.01.20143958v1; Makoto Kuroda, https://www.niid.go.jp/niid/images/research_info/genome-2020_SARS-CoV-MolecularEpidemiology_2.pdf (in Japanese).

[33] Nina Le Bert et al., Nature 584, 457-462 (2020) (https://doi.org/10.1038/s41586-020-2550-z); Annika Nelde et al., Research Square (2020), https://doi.org/10.21203/rs.3.rs-35331/v1; Takuya Sekine et al., Cell, in press (https://doi.org/10.1016/j.cell.2020.08.017).

[34] Iradj Amirlak et al., medRxiv (2020), https://doi.org/10.1101/2020.08.10.20172288; J. Sato, https://www.jsatonotes.com/.

